# Regular Non-steroidal Anti-inflammatory Drug Use Increases Stress Fracture Risk in the General Population: A Retrospective Case-Control Study

**DOI:** 10.1101/2023.10.30.23297788

**Authors:** Alexandra Ciuciu, Christopher Mulholland, Michael A. Bozzi, Chris C. Frymoyer, Leonardo Cavinatto, David Yaron, Marc I. Harwood, Jeremy D. Close, Christopher J. Mehallo, Ryan E. Tomlinson

## Abstract

Previous studies have shown that the use of non-steroidal anti-inflammatory drugs (NSAIDs) is associated with increased stress fracture risk, potentially due to diminished skeletal adaptation. This phenomenon has been studied predominantly in high-activity individuals like dancers, athletes, and soldiers, so current data regarding the general population is limited despite the substantial economic and resource burden of stress fracture injuries within the general US population. Furthermore, our preclinical studies demonstrate that regular use of NSAIDs also diminishes the intrinsic ability of bone to resist fracture. To determine the association of regular NSAID use with stress fractures in the general population, we surveyed subjects presenting with either stress fracture or uncomplicated ankle sprain to assess their use of NSAIDs over the three months before their injury. We hypothesized that subjects with stress fractures would have increased regular NSAID usage as compared to controls. Subjects diagnosed with a stress fracture (n=56) and subjects with uncomplicated ankle sprains (n=51; control) were surveyed about their NSAID use at the time of their diagnosis and in the previous three months using a questionnaire based on the National Health and Nutrition Examination Survey (NHANES). Subjects were surveyed in person on the day of their injury diagnosis or by phone within 30 days of their injury diagnosis. Fisher’s exact test was used to determine significant differences in NSAID usage between stress fracture and control subjects. Subjects diagnosed with stress fractures had a statistically significant increase in both current use (p=0.03) and regular use (p=0.04) of ibuprofen/naproxen/celecoxib as compared to control subjects. There were no significant differences in the use of aspirin, acetaminophen, or prescription medications containing acetaminophen between groups. Consistent with previous clinical reports, we observed a strong correlation between regular ibuprofen/naproxen/celecoxib use and stress fracture incidence in the general population. These results indicate that any patients at high risk for stress fracture should avoid regular use of ibuprofen, naproxen, or celecoxib.

**KEY POINTS:**

- This retrospective case-control study found that use of ibuprofen, naproxen, and/or celecoxib by subjects within the general population three or more times per week for three months (regular use) was highly associated with stress fracture diagnosis.
- No significant correlation between stress fracture diagnosis and the use of acetaminophen, aspirin, or prescription drugs containing acetaminophen was found.
- Pooled analysis of all surveyed drugs indicated a strong association between stress fracture diagnosis and NSAID or acetaminophen usage in the general population, consistent with previous work done in US Army soldiers and athletes.
- Due to the differences in mechanisms of action of the drugs surveyed in this study, future work is needed to conclude which NSAID, if any, is safest for regular use by those at higher risk of stress fracture formation.
- The findings of this study highlight the need for future studies to include subjects from the general population in addition to high-activity subjects as they indicate that these individuals are also at higher risk of stress fracture formation with regular NSAID use.

## INTRODUCTION

Non-steroidal anti-inflammatory drugs (NSAIDs) are common medications used to relieve pain, fever, and inflammation. Globally, NSAIDs are used by more than 30 million people daily and are often overused^1^. There is a growing body of pre-clinical evidence indicating that regular NSAID use diminishes bone formation in response to mechanical forces in areas of high strain, reduces bone toughness (the work required to fracture a bone), and impairs fracture healing^2–7^. Despite these observed negative effects of NSAIDs in bone, our understanding of the mechanisms behind the effects and which drugs cause them is incomplete. NSAIDs function by inhibiting the cyclooxygenase (COX) enzymes COX1 and/or COX2 from producing prostaglandins that can trigger pain or inflammation, but certain prostaglandins, particularly prostaglandin E2 (PGE2), are involved in strain adaptive bone remodeling and fracture healing^5,6,8^.

Strain adaptive bone remodeling and microdamage repair are bone functions that work in tandem to prevent stress fractures, which are painful fatigue injuries commonly caused by repetitive, submaximal forces on bone that from an osseous stress response that can propagate into larger fractures^9,10^. Stress fractures are commonly diagnosed in active individuals, such as athletes, dancers, and military personnel, and these injuries represent up to 20% of all injuries diagnosed in sports medicine clinics^2,9–11^. The treatment for a stress fracture often involves protected weight bearing and immobilization and can even require surgery. This imposes a large burden on the individual and the healthcare system. As a result, determining the factors that increase stress fracture incidence is a critical research goal.

Two recent studies have provided some mechanistic insights regarding the relationship of NSAIDs and stress fractures. First, one study assessed NSAID usage in US Army Soldiers^2^. Here, the investigators observed a 2.9-fold increase in stress fracture incidence associated with NSAID prescription in the general army population and a 5-fold increase in incidence for those in basic combat training. These results imply a general risk of NSAIDs in this active population that is increased during scenarios of particularly strenuous physical activity. Next, our laboratory investigated the impact of these NSAIDs in a preclinical mouse model^6^. Here, we observed a significant impact of the NSAID naproxen to decrease load-induced bone formation, as observed with NSAID administration in previous studies, as well as a novel deleterious effect on bone toughness^3,5,8,12,13^. We expect that both effects would increase stress fracture incidence, particularly when taken regularly.

Unfortunately, most studies on stress fracture incidence have been limited to athletes or military personnel, rather than the general population^9,10^. Therefore, the objective of this study was to quantify the association between regular NSAID use and stress fracture incidence in the general population. To do so, we used a retrospective case-control study of subjects diagnosed with a stress fracture as compared to subjects diagnosed with an uncomplicated ankle sprain (control). Our overall hypothesis was that subjects presenting with a stress fracture would have significantly higher regular NSAID use as compared to control subjects.

## METHODS

### Recruitment

This study protocol was approved by the IRB of Thomas Jefferson University (IRB #17D.318). Male and female participants (age range 18-83 years) recruited for this study between February and May of 2018 were diagnosed in our clinic with a stress fracture that was confirmed using MRI without contrast (ICD10 codes M84.30-M84.38). A total of 125 subjects diagnosed with stress fractures were considered, of which 69 subjects declined to participate. Subjects diagnosed with uncomplicated ankle sprains (ICD10 code S93.4) were used as controls and recruited between October of 2020 and February of 2021. A total of 214 ankle sprain subjects were considered, of which 163 declined to participate and 1 was excluded due to a preexisting condition causing chronic pain and frequent NSAID use. Overall, 56 subjects with stress fractures and 51 subjects with uncomplicated ankle sprains were recruited to participate in the study (Supplementary Figure 1).

### Questionnaire

To participate in this study, subjects were asked to complete a questionnaire in person or over the phone within 30 days of their injury diagnosis. First, each subject provided background information about their gender, age, height, weight, zip code, race, and the location of their stress fracture (if applicable). Next, their current and regular use of NSAIDs was determined using a series of questions taken from the National Health and Nutrition Examination Survey (NHANES)^14^. Specifically, we collected the following variables: ASPIRIN, ASPIRIN3M, ADVIL, ADVIL3M, ACETOCT, ACETOCT3M, ACETPR, and ACETPR3M (Table 1). Note that regular use is defined as three or more times per week for the previous three months.

**Table 1.** Medication variables. These variables were previously defined in the NHANES questionnaire.

| Drug Category | Included drugs |
| --- | --- |
| ASPIRIN, ASPIR3M | Aspirin, Bayer, Bufferin, Excedrin |
| ADVIL, ADVIL3M | Advil, Ibuprofen, Motrin, Nuprin, Aleve, Naprosyn, Naproxen, Celebrex, Celecoxib |
| ACETOCT, ACETOCT3M | Tylenol, Tylenol PM, Nyquil, Theraflu, Excedrin, Alka Seltzer Plus, Midol, acetaminophen |
| ACETPR, ACETPR3M | Vicodin, Percocet, Endocet, Tylenol with Codeine, Fioricet |

### Statistical analysis

Before analysis, data were de-identified and the results were analyzed for current and regular drug use between stress fracture and control subjects. Statistical analysis was performed using Fisher’s exact test in GraphPad Version 8.0.0. Resulting p-values less than 0.05 were considered significant.

## RESULTS

### Subject Demographics

Of the 56 stress fracture subjects and 51 uncomplicated ankle sprain (control) subjects screened for enrollment, one control subject was excluded due to chronic pain that resulted in regular NSAID use. With regards to cohort characteristics (Table 2), there were no significant differences between groups in mean age, sex, or race, with a small but significant difference in body mass index (BMI). Although not significantly different between groups, subjects in both cases and controls were predominantly female and white. The location of stress fractures in this cohort (Table 3) was primarily in the pelvis/femur (32%), the tibia/fibula (32%), and the ankle/foot/toe (30%).

**Table 2.** Subject Demographics. The demographic distribution of subjects within each injury category. Fisher’s exact test (sex), unpaired Student’s t-test (age, BMI), or Chi-square test for trend (race) were used to determine p-values. * p < 0.05, ns = not significant.

| Characteristic |  | Stress Fracture, n = 56 | Ankle Sprain (Control), n = 50 | P-value | Significance |
| --- | --- | --- | --- | --- | --- |
| Female sex - % |  | 73% | 64% | 0.40 | ns |
| Mean age (SD) - years |  | 51.4 (19.5) | 51.2 (13.9) | 0.96 | ns |
| Mean BMI (SD) – kg/m<sup>2</sup> |  | 27.1 (5.5) | 30.5 (8.7) | 0.02 | * |
| Race | White | 51 | 41 | 0.53 | ns |
|  | Black | 2 | 3 |  |  |
|  | Asian | 0 | 2 |  |  |
|  | Hispanic | 0 | 2 |  |  |
|  | Other | 0 | 1 |  |  |
|  | Prefer not to answer | 3 | 1 |  |  |

**Table 3.** Stress Fracture Location. The reported locations of stress fractures of subjects in the stress fracture injury group.

| Location of Stress Fracture | Stress Fracture, n | ICD-10 Code |
| --- | --- | --- |
| Shoulder | 1 | M84.31 |
| Humerus | 1 | M84.32 |
| Ulna or Radius | 0 | M84.33 |
| Hand or Finger | 0 | M84.34 |
| Pelvis or Femur | 18 | M84.35 |
| Tibia or Fibula | 18 | M84.36 |
| Ankle, Foot, or Toe | 17 | M84.37 |
| Other | 1 | M84.38 |

### Current NSAID use

First, we examined NSAID use at time of presentation with injury (Table 4). Here we observed that subjects with stress fractures had a significantly higher incidence of current use of drugs within the ADVIL category (p=0.025) as compared to control subjects (OR: 2.7, 95% CI: 1.1-6.1) (Figure 1). There were no significant differences between the percentage of subjects in the ASPIRIN, ACETOCT, or ACETPR groups. Nonetheless, we observed that subjects with stress fractures had significantly higher current use of any queried drug (p<0.0001) as compared to control subjects (OR: 8.5, 95% CI: 3.5-19) (Figure 2). Comparison of stress fracture subjects with subjects of the 2015 NHANES survey revealed that stress fracture subjects had significantly higher current use of drugs within the ADVIL and ACETOCT categories (Supplementary Table 1).

**Figure 1:**
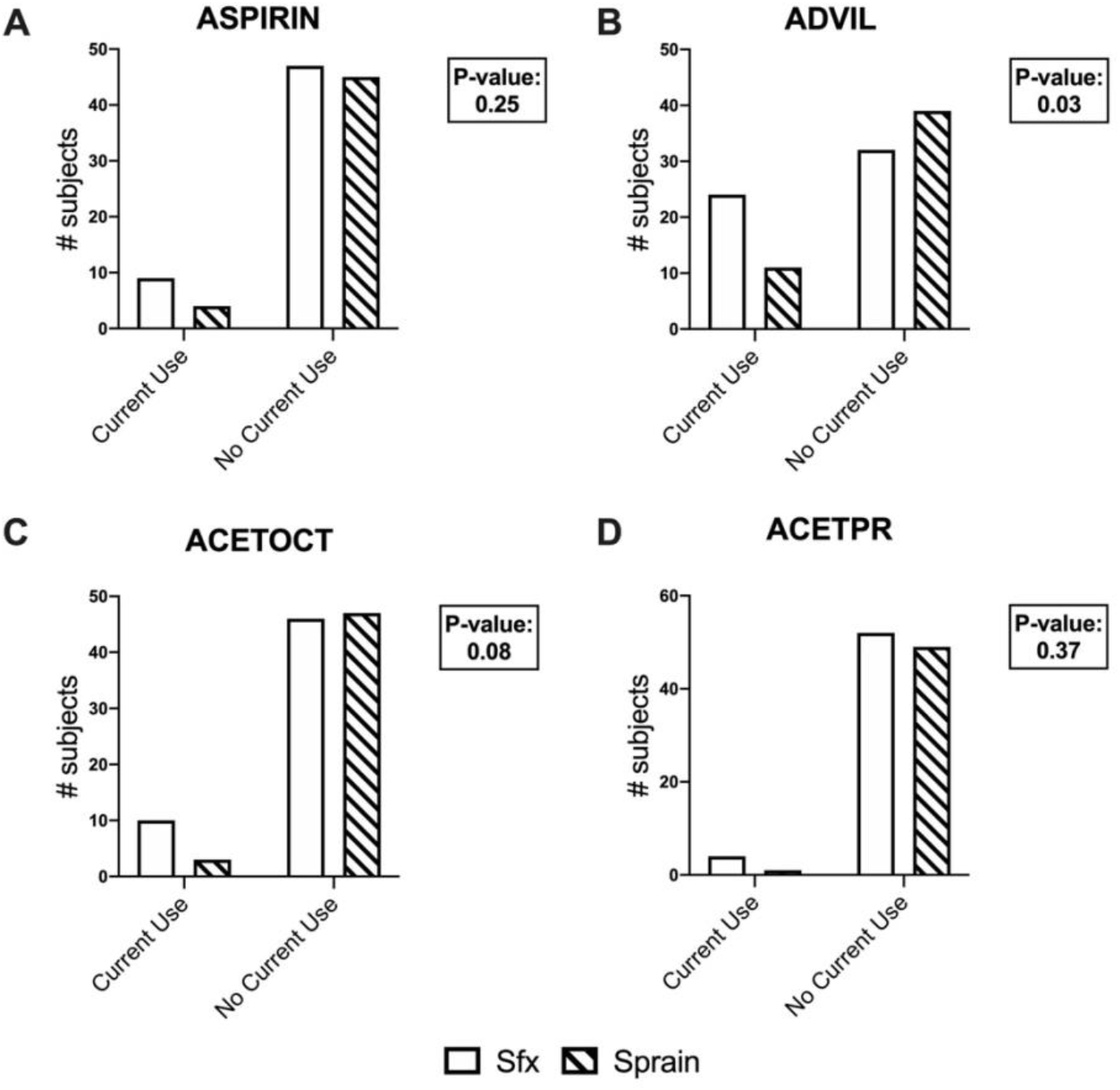
Current Drug Use by Category. Subjects diagnosed with stress fractures or uncomplicated ankle sprains answered Yes or No to survey questions regarding their current analgesic drug use, defined as 3 or more times per week. Results were divided into A) ASPIRIN, B) ADVIL, C) ACETOCT, and D) ACETPR. There was a significant increase in the amount of stress fracture subjects currently using drugs within the ADVIL category when compared to control subjects.

**Table 4.** Current Drug use. The enumerated responses of subjects within each injury group regarding current drug use defined as three or more times per week at the time of inquiry. * p < 0.05, ns = not significant.

| Drug Category | Stress Fracture, n |  |  | Ankle Sprain (Control), n |  |  | P-value | Significance |
| --- | --- | --- | --- | --- | --- | --- | --- | --- |
|  | YES | NO | NOT SURE | YES | NO | NOT SURE |  |  |
| **ASPIRIN** | 9 | 47 | 0 | 4 | 45 | 1 | 0.25 | ns |
| **ADVIL** | 24 | 32 | 0 | 11 | 39 | 0 | 0.03 | * |
| **ACETOCT** | 10 | 46 | 0 | 3 | 47 | 0 | 0.08 | ns |
| **ACETPR** | 4 | 52 | 0 | 1 | 49 | 0 | 0.37 | ns |
| **ALL Drug Categories** | 47 | 9 | 0 | 19 | 30 | 1 | <0.0001 | **** |

**Figure 2:**
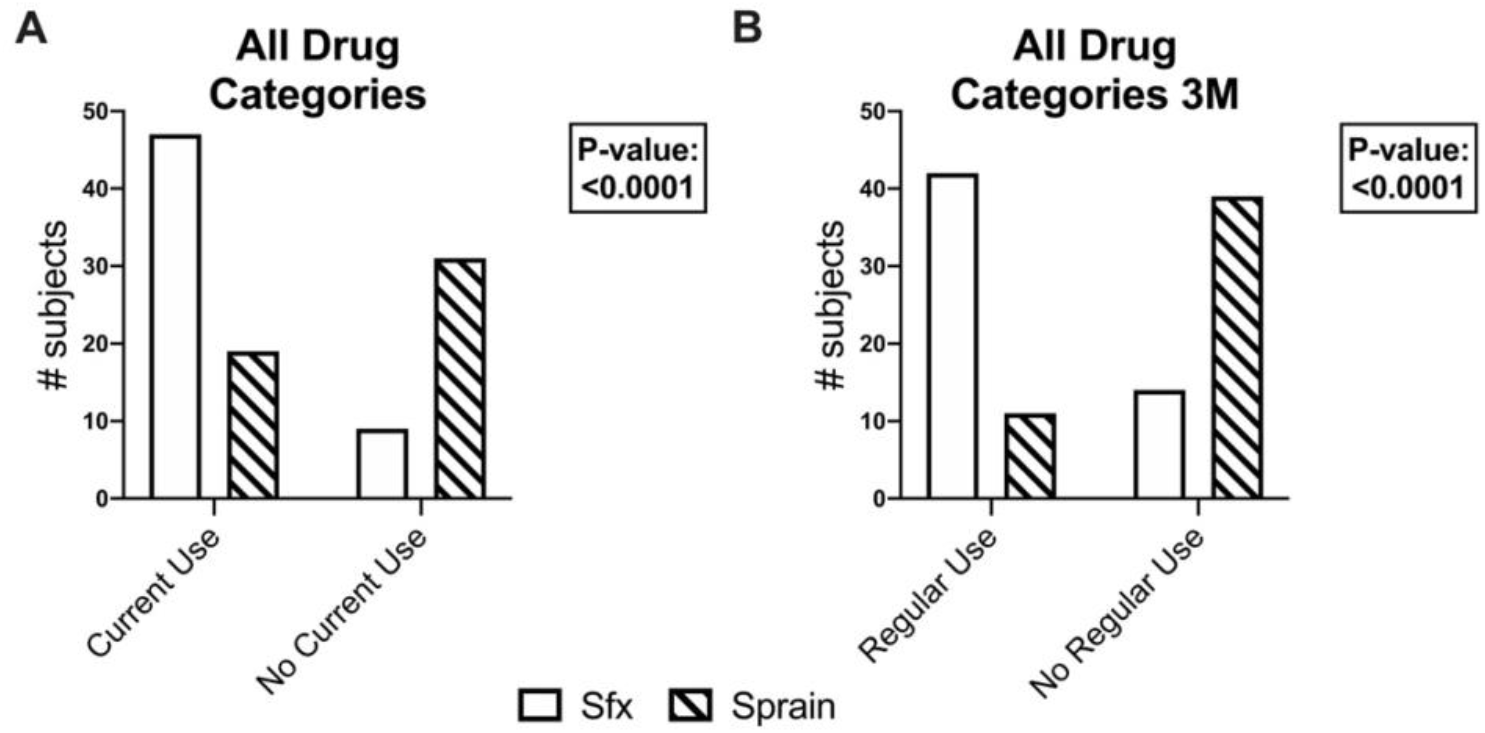
Current and Regular Use of All Surveyed Drugs. A) Total subject responses were pooled to quantify the number of stress fracture subjects compared to controls currently using any of the surveyed drugs. A significantly higher amount of stress fracture subjects currently used any of the surveyed drugs when compared to control subjects. B) Total subject responses were pooled to quantify the number of stress fracture subjects compared to controls using any of the surveyed drugs regularly. A significantly higher amount of stress fracture subjects regularly used any of the surveyed drugs when compared to control subjects. Overall, the use of any NSAID or acetaminophen-containing drug included in this surveyed was correlated to stress fracture diagnosis.

### Regular NSAID use

Next, we considered NSAID use in the previous three months before presentation with injury (Table 5), in which regular NSAID use was defined as three or more times per week during the three-month period. Here again, stress fracture subjects had a significantly higher incidence of regular use of drugs within the ADVIL category (p=0.041) as compared to the control subjects (OR: 6.0, 95% CI: 1.2 -26) (Figure 3). There were no significant differences between subjects in stress fracture and control groups for ASPIR3M, ACETOCT3M, or ACETPR3M. Finally, we analyzed pooled responses for regular use of any NSAID. We observed that stress fracture subjects had a significantly higher incidence of regular use of any queried drug (p<0.0001) as compared to control subjects (OR: 10.6, 95% CI: 4.1-25) (Figure 2). Comparison of stress fracture subjects with subjects of the 2015 NHANES survey revealed that there were no significant differences in regular NSAID use, potentially due to the inclusion of subjects with conditions of chronic pain (Supplementary Table 2).

**Table 5.** Regular Drug use. The enumerated responses of subjects within each injury group regarding regular drug use defined as three or more times per week for the three months prior to the time of inquiry. * p < 0.05, ns = not significant.

| Drug Category | Stress Fracture, n |  |  | Ankle Sprain (Control), n |  |  | P-value | Significance |
| --- | --- | --- | --- | --- | --- | --- | --- | --- |
|  | YES | NO | NOT SURE | YES | NO | NOT SURE |  |  |
| **ASPIRIN3M** | 9 | 0 | 0 | 4 | 0 | 0 | >0.99 | ns |
| **ADVIL3M** | 20 | 4 | 0 | 5 | 6 | 0 | 0.04 | * |
| **ACETOCT3M** | 9 | 1 | 0 | 1 | 2 | 0 | 0.11 | ns |
| **ACETPR3M** | 4 | 0 | 0 | 1 | 0 | 0 | >0.99 | ns |
| **ALL Drug Categories 3M** | 42 | 14 | 0 | 11 | 39 | 0 | <0.0001 | **** |

**Figure 3:**
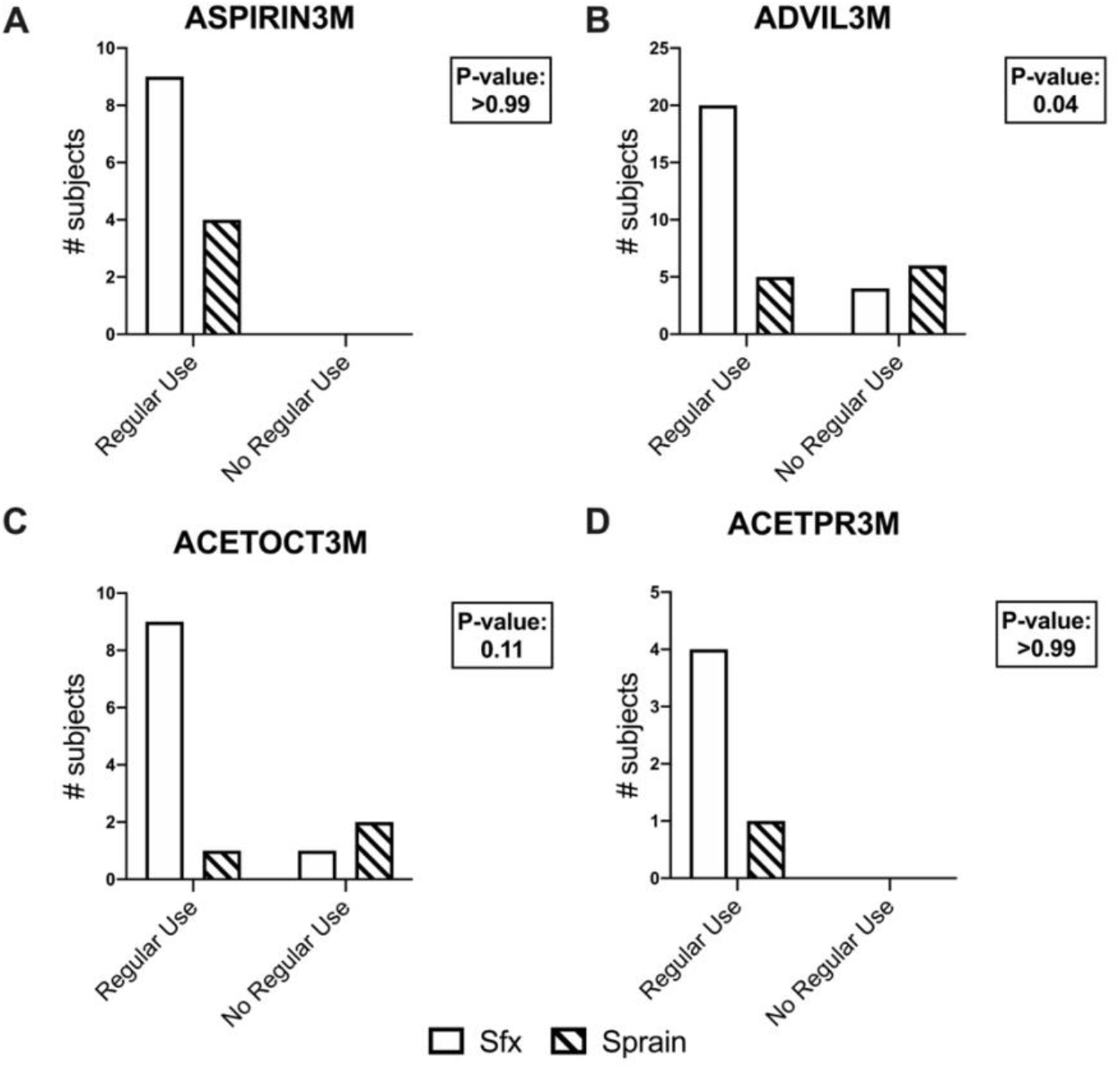
Regular Drug Use by Category. Subjects diagnosed with stress fractures or uncomplicated ankle sprains answered Yes or No to survey questions regarding their regular analgesic drug use, defined as 3 or more times per week for the past 3 months. Results were divided into A) ASPIRIN3M, B) ADVIL3M, C) ACETOCT3M, and D) ACETPR3M. There was a significant increase in the amount of stress fracture subjects currently using NSAIDs within the ADVIL3M category when compared to control subjects.

## DISCUSSION

In this study, we examined the relationship between regular NSAID use and stress fracture incidence. To do so, we assayed subjects with stress fracture or uncomplicated ankle sprain using the NHANES questionnaire. We found that regular use of ibuprofen, naproxen, and/or celecoxib was highly associated with stress fracture diagnosis. In contrast, we found no significant correlations between stress fracture diagnosis and the use of drugs containing acetaminophen, aspirin, or prescription drugs containing acetaminophen. Nonetheless, the pooled analysis of all drugs surveyed did indicate a strong association between stress fracture diagnosis and NSAID usage. In total, these results are consistent with our overall hypothesis and suggest that NSAIDs should not be used regularly by individuals engaged in activities associated with a high risk of skeletal injury.

This study contributes to a growing body of evidence that NSAIDs are associated with increased stress fracture risk ^5^. Importantly, these results from the general population are consistent with previous findings from a larger retrospective study of US Army Soldiers, in which naproxen and ibuprofen were associated with the highest stress fracture incidence ^2^. In addition to being the most popular NSAID category in our cohort, the drugs within the ADVIL category are potent COX2 inhibitors. As a result, subjects taking the ADVIL NSAIDs are likely to have decreased synthesis of prostaglandins, which have critical established roles in both strain adaptive bone remodeling and fracture healing ^3,5,6,8^. However, we cannot conclude that any one of the drugs in the ADVIL variable (ibuprofen, naproxen, celecoxib) is necessarily the most deleterious NSAID for bone health. In fact, our study reports that the odds for a subject with a stress fracture to have been taking any NSAID regularly (OR: 10.6, 95% CI: 4.1-25) is not significantly different than the odds for a subject with stress fracture to have been taking just the ADVIL NSAIDs regularly (OR: 6.0, 95% CI: 1.2-26). Since ibuprofen, naproxen, and celecoxib have substantially different COX selectivity, typical dosages, and strength of COX inhibition, these NSAIDs should be evaluated separately in future experimental work.

Our study did not report any significant correlation between current or regular use of aspirin, acetaminophen, or prescription medications containing acetaminophen and stress fracture incidence. This lack of correlation could be due to the action of these NSAIDs. Aspirin causes irreversible inhibition of the COX enzymes and the formation of several metabolic byproducts that are produced quickly after ingestion, which may limit its effects in bone compared to other NSAIDs ^15^. Contrastingly, acetaminophen is not an NSAID, but its analgesic and antipyretic actions in humans are believed to be mediated through binding with the COX enzymes. Also, acetaminophen may inhibit strain adaptive bone remodeling and fracture healing similarly to NSAIDs ^15,16^. Finally, the prescription medications containing acetaminophen are less accessible than over the counter analgesic medications and had the least number of users in both injury categories. Despite these contributing factors, the effects of these medications should continue to be investigated to minimize stress fracture risk in the general population. In particular, previous studies have demonstrated effects of NSAIDs and acetaminophen that are independent of COX inhibition, so directly assessing the effects of these medications on bone overuse injuries, rather than COX-dependent signaling, is required ^17,18^.

Here, we performed a retrospective, case-control study using subjects with uncomplicated ankle sprains as our control group. These individuals, like subjects with stress fractures, are generally mobile and present with a painful musculoskeletal injury. However, we also used the results of the 2015 NHANES survey as an additional control group to compare both our stress fracture and uncomplicated ankle sprain cohorts to an uninjured cohort (Supplementary Tables 1-4). Although our stress fracture cohort displayed significantly more current use of NSAIDs than the 2015 NHANES cohort, there were no differences in regular usage between groups. In contrast, we observed that subjects within the uncomplicated ankle sprain cohort displayed less regular use of NSAIDs— particularly those within the ADVIL category—as compared to subjects in the 2015 NHANES cohort. As a result, future studies done in the general population may need to employ matching between cases and controls as well as additional covariates (e.g. chronic disease) to control for the use of NSAIDs outside of musculoskeletal pain.

This study has several limitations, including the relatively small subject cohort that was predominantly female, white, and older—the population most at risk for osteopenia and osteoporosis. Since the effects of NSAIDs may be sex, race, and age dependent, this may limit the application of our findings ^19^. Moreover, the physical activity level of the subjects was not assayed. In addition to the central role of physical activity on stress fracture incidence, previous studies have shown that administration of NSAIDs before or after exercise affects the skeletal consequences ^3,5,12,13^. However, the only significant difference between stress fracture and control subjects was BMI, with the ankle sprain cohort having a slightly higher average BMI (30.5 kg/m^2^) than the stress fracture cohort (27.1 kg/m^2^) – perhaps indicating an overall higher activity level in the stress fracture subjects ^20^. Importantly, it is not known whether subjects in the stress fracture cohort developed their injury before or during the three-month period preceding their clinical presentation. To limit potential recall bias, subjects were asked about their NSAID usage as soon after their diagnosis as possible. A final limitation is that a stress fracture is an overuse injury that often takes longer to form and diagnose than an ankle sprain, which is often due to a single traumatic event. This fact, coupled with the regular access to over-the-counter drugs including ibuprofen, naproxen, and acetaminophen means that it is possible that subjects were self-medicating with the surveyed drugs before their official diagnosis. Additionally, subjects have more access to over-the-counter medications than prescribed drugs, which may have led to the diminished sample sizes within the ACETPR group. Future studies should consider drug accessibility as a factor when recruiting subjects. Despite these limitations, the overall study conclusions are consistent with previous clinical and preclinical studies regarding the strong correlation between NSAIDs and stress fracture risk.

## CONCLUSION

Our study found a significant correlation between the regular use of ibuprofen, naproxen, and/or celecoxib with stress fracture diagnosis in the general population. This finding is consistent with preclinical studies in animal models as well as clinical data from US Army Soldiers^2,5,6,11^. NSAID use likely increases the risk of stress fracture by decreasing inherent toughness and diminishing the ability of bone to repair microdamage and remodel in response to mechanical loading. Future studies should include observation of a more diverse study population as well as mechanistic experiments to investigate the effect of individual NSAIDs. Nonetheless, this study is one of a growing body of literature that has linked stress fracture incidence with NSAID usage. As a result, it is recommended that patients at high risk for stress fracture refrain from regular NSAID usage.

## Data Availability

All data generated or analyzed in this study are included in this published article and its supplemental data file.

