## Supplemental Data for "Regular Non-steroidal Anti-inflammatory Drug Use Increases Stress Fracture Risk in the General Population: A Retrospective Case-Control Study"

| **Drug Category** | **Stress Fracture, n** | | | **2015 NHANES, n** | | | **P-value** | **Significance** |
| --- | --- | --- | --- | --- | --- | --- | --- | --- |
|  | YES | NO | NOT SURE | YES | NO | NOT SURE |  |  |
| ASPIRIN | 9 | 47 | 0 | 6773 | 24529 | 29 | 0.42 | ns |
| ADVIL | 24 | 32 | 0 | 5404 | 25880 | 35 | <0.0001 | ******* |
| ACETOCT | 10 | 46 | 0 | 2915 | 28353 | 44 | 0.04 | * |
| ACETPR | 4 | 52 | 0 | 1257 | 228 | 56 | 0.34 | ns |

**Supplementary Table 1.** Current Drug Use. Stress fracture subjects compared to 2015 NHANES regions 1-4 questionnaire responses for current drug use defined as three or more times per week at time of questionnaire response. * Indicates a significant p value of $\leq$0.05. *** Indicates a highly significant p value of $\leq$0.001.

| **Drug Category** | **Stress Fracture, n** | | | **2015 NHANES, n** | | | **P-value** | **Significance** |
| --- | --- | --- | --- | --- | --- | --- | --- | --- |
|  | YES | NO | NOT SURE | YES | NO | NOT SURE |  |  |
| ASPIR3M | 9 | 0 | 0 | 6250 | 518 | 3 | >0.99 | ns |
| ADVIL3M | 20 | 4 | 0 | 4187 | 1213 | 3 | 0.63 | ns |
| ACETOC3M | 9 | 1 | 0 | 2128 | 786 | 1 | 0.31 | ns |
| ACETPR3M | 4 | 0 | 0 | 1257 | 228 | 0 | >0.99 | ns |

**Supplementary Table 2.** Regular Drug Use. Stress fracture subjects compared to 2015 NHANES regions 1-4 questionnaire responses for regular drug use defined as three or more times per week for three months prior to questionnaire response.

| **Drug Category** | **Uncomplicated Ankle Sprain, n** | | | **2015 NHANES, n** | | | **P-value** | **Significance** |
| --- | --- | --- | --- | --- | --- | --- | --- | --- |
|  | YES | NO | NOT SURE | YES | NO | NOT SURE |  |  |
| ASPIRIN | 4 | 45 | 1 | 6773 | 24529 | 29 | 0.022 | * |
| ADVIL | 11 | 39 | 0 | 5404 | 25880 | 35 | 0.35 | ns |
| ACETOCT | 3 | 47 | 0 | 2915 | 28353 | 44 | 0.62 | ns |
| ACETPR | 1 | 49 | 0 | 1257 | 228 | 56 | >0.99 | ns |

**Supplementary Table 3.** Current Drug Use. Uncomplicated ankle sprain subjects compared to 2015 NHANES regions 1-4 questionnaire responses for regular drug use defined as three or more times per week at time of questionnaire response. * Indicates a significant p value of $\leq$0.05.

| **Drug Category** | **Uncomplicated Ankle Sprain, n** | | | **2015 NHANES, n** | | | **P-value** | **Significance** |
| --- | --- | --- | --- | --- | --- | --- | --- | --- |
|  | YES | NO | NOT SURE | YES | NO | NOT SURE |  |  |
| ASPIR3M | 4 | 0 | 0 | 6250 | 518 | 3 | >0.99 | ns |
| ADVIL3M | 5 | 6 | 0 | 4187 | 1213 | 3 | 0.021 | * |
| ACETOC3M | 1 | 2 | 0 | 2128 | 786 | 1 | 0.18 | ns |
| ACETPR3M | 1 | 0 | 0 | 1257 | 228 | 0 | >0.99 | ns |

**Supplementary Table 4.** Regular Drug Use. Uncomplicated ankle sprain subjects compared to 2015 NHANES regions 1-4 questionnaire responses for regular drug use defined as three or more times per week for three months prior to questionnaire response.

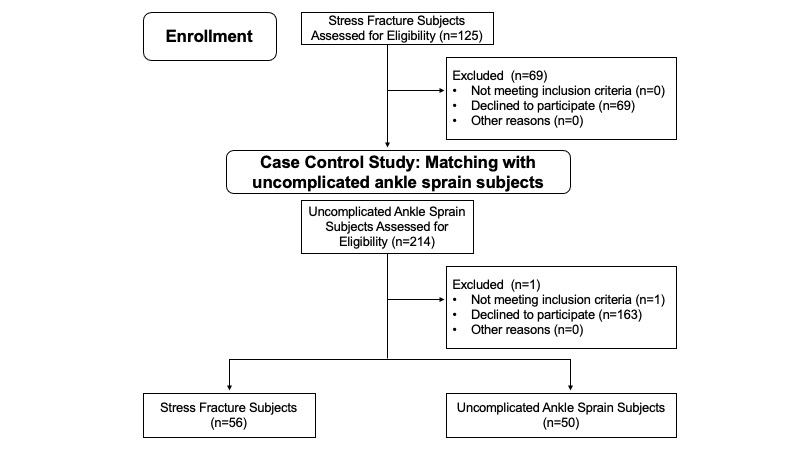

**Supplementary Figure 1.** CONSORT Flow Diagram. Subject selection process.
